# Genetic and clinical characteristics of treatment-resistant depression using primary care records in two UK cohorts

**DOI:** 10.1101/2020.08.24.20178715

**Authors:** Chiara Fabbri, Saskia P. Hagenaars, Catherine John, Alexander T. Williams, Nick Shrine, Louise Moles, Ken B. Hanscombe, Alessandro Serretti, David J. Shepherd, Robert S. Free, Louise V. Wain, Martin D. Tobin, Cathryn M. Lewis

## Abstract

Treatment-resistant depression (TRD) is a major contributor to the disability caused by major depressive disorder (MDD). Using primary care electronic health records from UK Biobank and EXCEED studies, we defined MDD and TRD, providing an easily accessible approach to investigate their clinical and genetic characteristics.

MDD defined from primary care records was compared with other measures of depression and validated using the MDD polygenic risk score (PRS). Using prescribing records, TRD was defined from at least two switches between antidepressant drugs, each prescribed for at least six weeks. Clinical-demographic characteristics, SNP-heritability and genetic overlap with psychiatric and non-psychiatric traits were compared in TRD and non-TRD MDD cases.

In 230,096 and 8,926 UKB and EXCEED participants with primary care data, respectively, the prevalence of MDD was 8.7% and 14.2%, of which 13.2% and 13.5% was TRD (2,430 and 159 cases), respectively. In both cohorts, MDD defined from primary care records was strongly associated with MDD PRS, and in UKB it showed overlap of 72%-88% with other MDD definitions. In UKB, TRD and non-TRD heritability was comparable (h^2^_SNP_ = 0.25 [SE=0.04] and 0.19 [SE=0.02], respectively). TRD was positively associated with the polygenic risk score (PRS) of attention deficit hyperactivity disorder and negatively associated with the PRS of intelligence compared to non-TRD. It was more strongly associated with unfavourable clinical-demographic variables than non-TRD.

This study demonstrated that MDD and TRD can be reliably defined using primary care records and provides the first large scale population assessment of the genetic, clinical and demographic characteristics of TRD.

## 1. Introduction

Major depressive disorder (MDD) is a common psychiatric disorder affecting more than 264 million people worldwide, and is the fourth-leading cause of disability (GBD 2015 Disease and Injury Incidence and Prevalence Collaborators, 2016; James et al., 2018), other than a major contributor to the death by suicide of one person every 40 seconds in the world (World Health Organization, 2019).

The study of factors determining MDD pathogenesis, prognosis and response to treatments has been a major research area, with the aim of providing better instruments for disease prevention, early diagnosis and personalized treatment, to facilitate recovery. Recovery, defined as complete remission of depressive symptoms, is associated with better functioning and reduced risk of depressive relapse (Gaynes et al., 2009). However, a substantial proportion of MDD patients do not reach remission after multiple antidepressant treatments. These patients are classified as having treatment-resistant depression (TRD), usually defined as lack of response to at least two antidepressant treatments. Using this definition, the prevalence of TRD in MDD cases was estimated as 7% in Scottish primary care (based on Electronic Health Records [EHR]) and 22% in Canada primary care (based on a questionnaire filled by physicians) (Wigmore et al., 2020; Rizvi et al., 2014). TRD is a particularly heavy burden for the patient and society, being associated with social and occupational impairment, suicidal thoughts, decline of physical health, increased health care utilization and higher all-cause mortality compared to non-TRD (Trivedi et al., 2006; Souery et al., 2011; Li et al., 2019).

Antidepressants are a first line treatment for MDD, with over 40 compounds currently available. Network meta-analysis shows clear benefit of antidepressants over placebo with some differences between drugs, but there are large inter-individual differences in response. In the UK, most MDD treatment occurs in primary care and antidepressant treatment is recommended for moderate to severe depression (Ferenchick et al., 2019). Non-response to antidepressants is common, with only 45% of UK primary care patients responding to their current antidepressant treatment taken for at least 6 weeks (Thomas et al., 2013).

Resistance to antidepressants is partly influenced by genetic factors, with a SNP-heritability of 0.132 (SE=0.056) for remission (Pain et al., 2019), while TRD was estimated to have a heritability of 0.60 (SE=0.19) from pedigree data (Wigmore et al., 2020). Two genome-wide association studies (GWAS) have identified no genetic variants associated with TRD (Wigmore et al., 2020; Fabbri et al., 2019). The largest TRD GWAS used self-reported data from 23andMe (N = 1,311 TRD, N = 7,795 non-TRD) (Li et al., 2016). This study did not identify any variants associated with TRD and reported a non-significant heritability of TRD vs non-TRD and a heritability of 17% (95% CI = 6% - 27%) for TRD vs healthy controls. Few studies have had sufficient power to investigate the genetic overlap of TRD with other psychiatric traits but available results suggest a genetic correlation of TRD with personality traits such as neuroticism and general health (Wigmore et al., 2020). No studies of genetic correlation with non-psychiatric traits have been reported.

Current studies in TRD have been hampered by small sample sizes and low power, but EHR provide an exciting opportunity for large scale pharmacogenetics research studies at low cost and with high classification accuracy (Smoller, 2018). Most diseases, including psychiatric disorders, are well coded in primary care records, with chronic diseases having a positive predictive value > 90% (Hardoon et al., 2013; Khan et al., 2010). Most patients with MDD are treated in primary care, and primary care records should be a valuable research resource to improve the clinical management of MDD in health care systems.

In this study, we used primary care EHR linked to the Extended Cohort for E-health, Environment and DNA (EXCEED) cohort to develop and test an algorithm to define MDD and TRD; then, this was applied to UK Biobank (UKB) to validate and extend the results to a larger cohort. Specifically, using the primary care data, we

1. Identified patients with MDD, validating our algorithm in EXCEED and UKB by assessing genetic overlap with an independent and large MDD sample, and through phenotypic comparisons with other MDD definitions available in UKB;
2. Identified patients with TRD, then studied their clinical and socio-demographic features (both cohorts), calculated TRD heritability and assessed genetic correlations with other psychiatric and non-psychiatric traits (UKB).

## 2. Materials and Methods

### 2.1. EXCEED and UK Biobank cohorts

The EXCEED cohort includes over 10,000 individuals from Leicester, Leicestershire and Rutland with genetic data and linkage to primary care and hospital EHR. Data includes longitudinal health information from EHR from primary care and Hospital Episode Statistics. Primary care data was available for 8,926 participants. Date and clinical code (Read v2 or CTV3) are available for primary care events (John et al., 2019). As EXCEED primary care EHR data were available before similar data in UKB, this extensively annotated, smaller data set was used to develop the algorithms to define MDD and TRD, and then for comparison of the results.

UKB is a prospective population-based study of ~500,000 individuals recruited across the United Kingdom, aged between 40-69 at baseline. To date, coded primary care data have been obtained for approximately 45% of the cohort (~230,000 participants, all of whom have provided written consent for linkage to their health-related records) (UK Biobank, 2019). Clinical code (Read v2 or CTV3) and associated dates are available for primary care events, such as consultations and diagnoses. Drug code (Read v2, BNF 2 and/or dm+d), associated dates and, where available, drug name and quantity, are reported for medicines prescribed in primary care (UK Biobank, 2019). Further information is available in Supplementary Methods.

### 2.2. Definition of MDD and TRD

In both cohorts, participants with MDD were identified as those having:

– At least two diagnostic codes for any unipolar depressive disorder, at any time point (two codes were required to reduce the risk of miscoding and inclusion of individuals with main diagnosis other than MDD);
– No diagnostic code for bipolar disorders, psychotic disorders or substance use-related disorders.

Participants with TRD were defined as those with MDD having at least two switches between different antidepressant drugs satisfying the following criteria (Figure 1):

**Figure 1:**
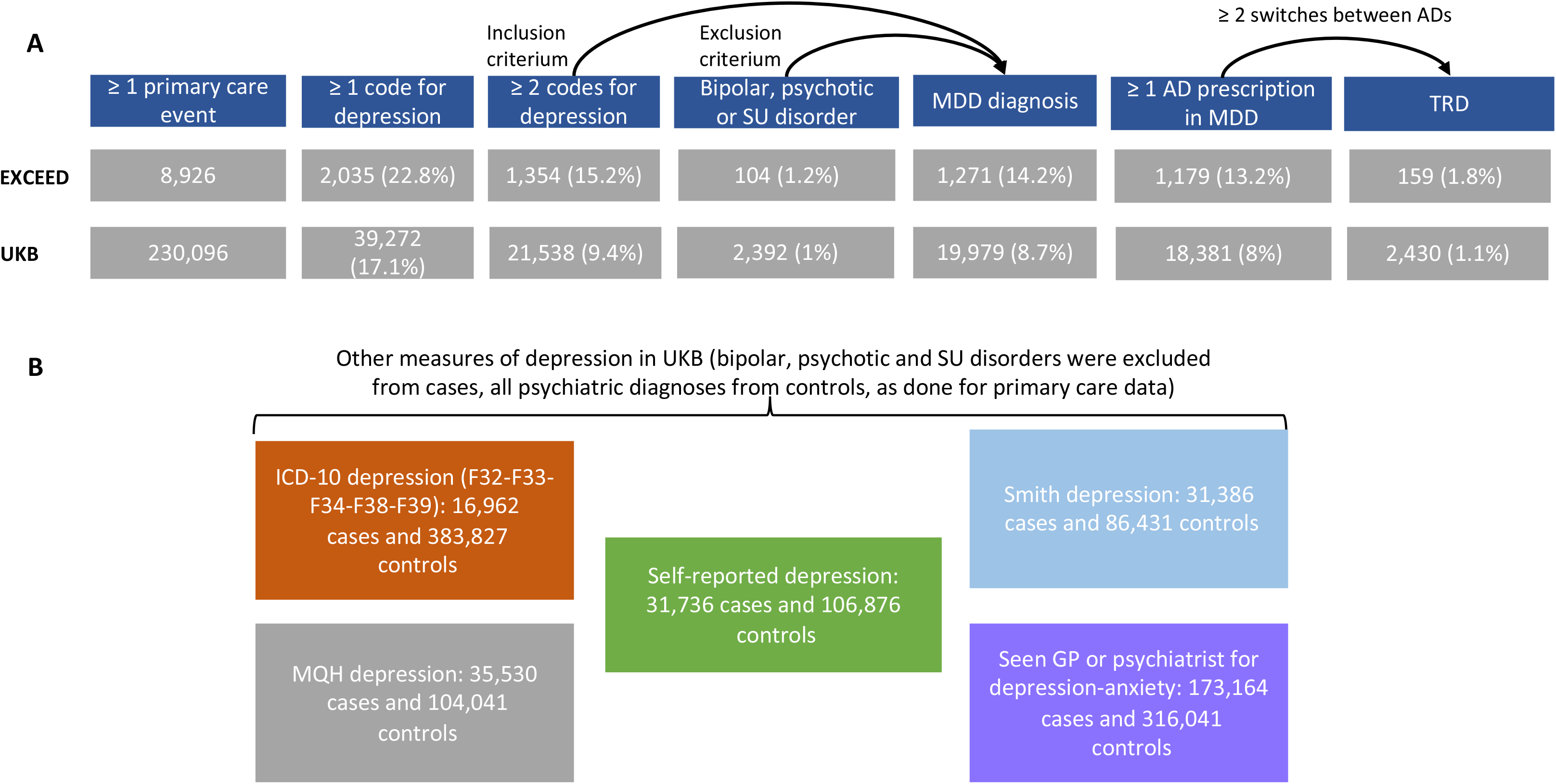
selection of individuals with depression and treatment-resistant depression (TRD) in EXCEED and UKB primary care data **(A)** and other measures of depression used as comparison in UKB **(B)**. MDD=major depressive disorder; SU=substance use; AD=antidepressant.

– Each drug was prescribed for at least 6 consecutive weeks (noting that adequate duration for efficacy is four weeks, and our conservative threshold should reduce the risk that drug switch was due to side effects, in line with a previous study (Wigmore et al., 2020));
– The time interval between two consecutive drugs was no longer than 14 weeks (to ensure that treatment had not been suspended and that the drugs were prescribed for the same episode of depression).

The codes corresponding to the primary care events (diagnoses and medications) used to define these phenotypes are in Supplementary Table 1 and the code used to extract them and define the phenotypes in UKB is Supplementary code 1.

To evaluate a proxy of poor compliance to treatment as a possible contributor to the poorer response observed in TRD, for each subject we calculated the proportion of adequate prescription intervals, defined by 14 weeks or less between subsequent antidepressant prescriptions.

### 2.3. Genotyping, quality control and imputation

To date, over 60% of EXCEED participants have been genotyped using the Affymetrix UK Biobank Axiom Array. Data were available in 5216 participants after quality control (see Supplementary Methods), with imputation to the Haplotype Reference Consortium (HRC) panel (McCarthy et al., 2016).

Genome-wide genotyping on all UK Biobank participants was performed using two highly overlapping arrays covering ~800,000 markers. The description of quality control is reported in Supplementary Methods. A two-stage imputation was performed using the Haplotype Reference Consortium (HRC) and UK10K reference panels (Bycroft et al., 2018; McCarthy et al., 2016; UK10K Consortium et al., 2015). Poor imputed variants were excluded (IMPUTE INFO metric <= 0.4) (McCarthy et al., 2016).

### 2.4. Statistical analyses

#### 2.4.1. Phenotypic analyses

To validate the definition of MDD using primary care EHR, we cross-classified with five UKB MDD phenotypes (Figure 1): (1) the Mental Health Questionnaire (MHQ) (Davis et al., 2020), (2) hospital diagnosis (ICD-10 codes F32-F33-F34-F38-F39) (data fields 41202 and 41204), (3) self-reported depression diagnosed by a professional (data field 20544), (4) help-seeking for depression (data fields 2090 and 2100) and (5) Smith et al. definition (Smith et al., 2013) (Supplementary Methods). In both cohorts, TRD can only be defined using primary care EHR, so we assessed the clinical and socio-demographic characteristics of TRD and non-TRD cases and we compared these findings with the existing literature on TRD. In UKB, we also tested if antidepressant combinations or augmentation with an antipsychotic or mood stabilizer were more common in TRD than non-TRD cases, as these pharmacological strategies are suggested by prescribing guidelines for TRD (Taylor et al., 2018). Only combination or augmentation prescriptions for over 30 days were included. A description of these medications, diagnostic codes of psychiatric comorbidities and the code used to extract them are in Supplementary Table 2 and Supplementary code 2, respectively (UKB).

#### 2.4.2. Polygenic risk scores, genetic correlations and heritability

We calculated polygenic risk scores (PRS) for MDD (Wray et al., 2018), schizophrenia (Pardiñas et al., 2018) and bipolar disorder (Stahl et al., 2019) to test their association with MDD in primary care and other depression phenotypes described above. We hypothesized that prediction would be stronger for MDD PRS than other psychiatric disorders (both cohorts). Participants with no psychiatric diagnoses based on primary care EHR were used as healthy controls; for each of the other depression phenotypes in UKB, those with no psychiatric diagnoses according to that measure were included as healthy controls (Figure 1).

PRS were calculated using PRSice v.2 (Choi and O’Reilly, 2019) and genotyped variants at 11 p-value thresholds *P_T_* (5e-8, 1e-5, 1e-3, 0.01, 0.05, 0.1, 0.2, 0.3, 0.4, 0.5, 1); the most predictive *P_T_* was selected (see Supplementary Methods for further details). Logistic regression models were used to estimate associations between the phenotype and each PRS adjusting for six genetic ancestry principal components, assessment centre and batch effects in UKB (Supplementary code 3) and six genetic ancestry principal components and primary care practice in EXCEED. The proportion of variance explained by PRS on the liability scale was estimated according to Lee et al. (Lee et al., 2012), assuming MDD prevalence of 10.8% for case-control comparisons (Lim et al., 2018). A Bonferroni correction was applied considering the number of traits and *P_T_* tested.

For the estimation of heritability (*h^2^_SNP_*) of TRD and non-TRD in UKB (these analyses would not have adequate power in EXCEED), we used genome-wide complex trait analysis software v.1.93.1beta (GCTA) (Yang et al., 2011) and genetic relationship matrix-restricted maximum likelihood (GREML). The genetic relationship matrix was adjusted for incomplete tagging of causal SNPs and we further excluded related individuals using a grm-cut off of 0.05 (Supplementary code 4). We included 11,188 healthy controls (no psychiatric diagnoses) as they provided adequate power assuming a prevalence of 0.02 of TRD in the population and heritability of 0.10 for MDD (Visscher et al., 2014; Wray et al., 2018).

We also calculated *h^2^_SNP_* using Genome-wide Complex Trait Bayesian (GCTB) Bayes S method. GCTB uses the data to estimate polygenicity (i.e., the proportion of SNPs with nonzero effects, *P_i_*), in contrast to GCTA-GREML which assumes that all SNPs have an effect on the trait. GCTB also calculates the relationship between effect size and MAF (5) which can be used to detect signatures of natural selection (Zeng et al., 2018). Heritability estimates from GCTA-GREML and GCTB were compared with results from linkage disequilibrium score regression (LDSC) (Zheng et al., 2017). For all analyses, six genetic ancestry principal components, assessment centre and batch effects were included as covariates (Supplementary code 4).

*h^2^_SNP_* was transformed to the liability scale using a range of possible population prevalences (Yap et al., 2018) (Lim et al., 2018) (Rizvi et al., 2014). We evaluated the possibility that heritability estimates may be inflated by selecting extremes of the controls (individuals without any psychiatric disorder) and cases (individuals with at least two diagnostic records of depression) distributions (Yap et al., 2018). This was performed by comparing *h^2^_snp_* obtained using a set of controls of the same size with no MDD but without screening for other possible psychiatric disorders and by considering the prevalence of MDD with two or more diagnostic records of depression among subjects with at least one diagnostic record of depression in UKB (Yap et al., 2018) (Supplementary code 4).

Genetic correlations (*r_g_*) with selected psychiatric and non-psychiatric traits were estimated using LDSC with GWAS summary statistics available on LD Hub (Zheng et al., 2017) (Supplementary Table 3). Three GWAS were performed: TRD vs. non-TRD, TRD vs. 11,188 healthy controls and non-TRD vs. 11,188 healthy controls (see above for the definition of healthy controls). GWAS were performed using BGENIE v1.2 and imputed genotype dosages (Bycroft et al., 2018), with phenotypes residualised for six genetic ancestry principal components, assessment centre and batch effects (Supplementary code 5).

*R_g_* estimates and previous studies on TRD were used to guide the selection of PRS tested for association with TRD vs. non-TRD (Wigmore et al., 2020). A list of all traits used for PRS analyses and the respective summary statistics is reported in Supplementary Table 4. Bonferroni correction was applied to take into account multiple testing.

## 3. Results

In EXCEED and UKB, 8,926 and 230,096 participants had at least one primary care event, and the prevalence of MDD was 14.24% (n=1,271) and 8.68% (n=19,979), respectively. In both cohorts, the majority of depression diagnoses were from 1990-2017, with a prevalence of MDD of 12.62% and 8.5% over this time frame in EXCEED and UKB, respectively. Among individuals with MDD with at least one record of antidepressant prescription, the prevalence of TRD was 13.49% (n=159) and 13.2% (n=2,430) in EXCEED and UKB, respectively (Figure 1).

In UKB we also looked at trends of depression diagnoses and antidepressant prescriptions (drugs and classes) over time, since UKB had larger sample size and was more representative than EXCEED (Supplementary Figure 1); to provide a comprehensive picture, these figures include all participants with at least one diagnostic code for depression but no code for bipolar disorders, psychotic disorders or substance use disorders (n=36,880). The increasing number of diagnoses and antidepressant prescriptions across time reflects the increasing completeness of primary care EHR (UK Biobank, 2019), but also captures real trends. For example, antidepressant prescriptions increased by 10.2% from 2003 to 2004, reflecting a general increase across time, but this flattened to an increase of only 2.6% in 2005, as a probable consequence of the “black box” warning on the risk of antidepressant-induced suicidality in 2004 (Stone, 2014).

### 3.1. Validation of primary care diagnosed MDD

#### 3.1.1. PRS of MDD, schizophrenia and bipolar disorder

In the EXCEED and UKB cohorts, 557 and 17,807 participants with MDD had genetic data after quality control, and 2,181 and 130,356 controls with no psychiatric diagnosis, respectively. MDD PRS was associated with primary care-defined MDD diagnosis (beta=0.23 (SE=0.05), p=6.05e-06 and beta=0.15 (SE=0.008), p=2.73e-71, in EXCEED and UKB, respectively), with a similar effect size in the two cohorts (z test to compare beta and SE of MDD PRS in the two samples: z=1.58, p=0.11). In EXCEED, schizophrenia and bipolar disorder PRS was not associated with MDD case-control status after multiple-testing correction (Supplementary Table 5), while in UKB they had a significant effect (because of higher power in this sample) that however was smaller than the effect of MDD PRS (z=5.03, p=4.91e-07 and z=6.41, p=1.46e-10 for schizophrenia and bipolar disorder PRS, respectively). The Nagelkerke *R^2^* (liability scale) of MDD PRS was 1.2% and 0.6% in EXCEED and UKB, respectively (Supplementary Table 5).

Given the larger sample size and greater generalizability of results, in UKB we compared the association of MDD PRS with depression defined using at least one vs. at least two diagnostic codes for depression and found no difference (z=0.81, p=0.42). Both these definitions showed similar associations with MDD PRS when compared to other measures of depression in UKB (Figure 2, Supplementary Table 5).

**Figure 2:**
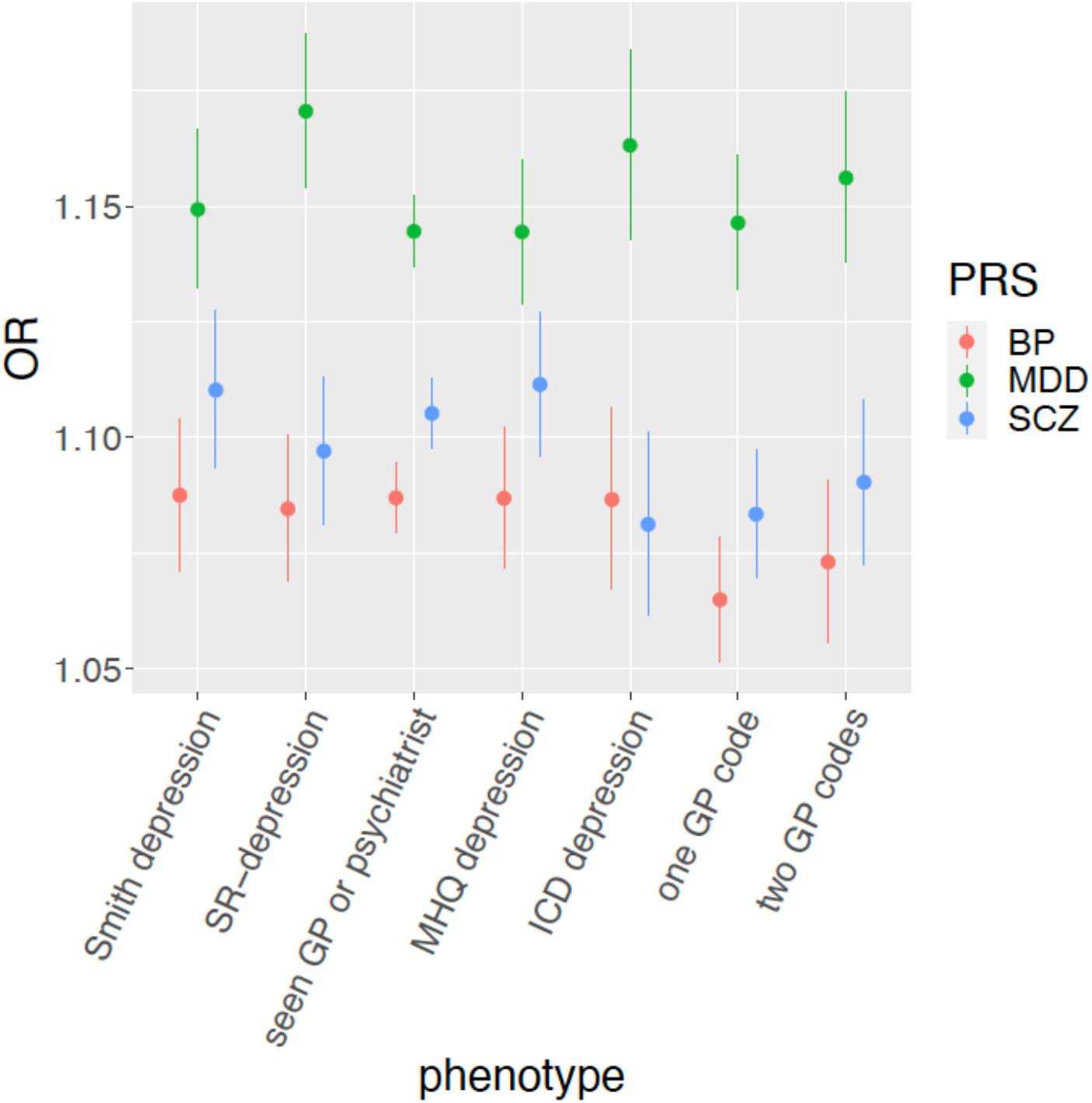
Association between polygenic risk scores (PRS) for psychiatric disorders and depression phenotypes in UKB, showing odds ratios (OR) and 95% confidence intervals. SR-depression=self-reported depression diagnosed by a professional; MHQ=mental health questionnaire; GP=general practitioner.

#### 3.1.2. Comparison with other depression measures in UKB

MDD defined from primary care data showed overlap with other measures of depression in 72%-88% of cases in UKB; 20% of participants with MDD according to primary care records also received a diagnosis of depression in a hospital setting (ICD-10 codes), while 72% of patients with an ICD-10-code for a depressive disorder had at least one diagnostic code for depression in primary care data (93% of these were ICD-10 codes for secondary diagnoses). Individuals with an ICD-10-code for depression who had at least one diagnostic code for depression in primary care data had an increased probability that their ICD-10-code was a main diagnosis code rather than a secondary diagnosis code (OR=1.51, [1.19-1.92], p=3.76e-04), suggesting that secondary diagnoses may not have been followed up in primary care. For all the considered measures except MHQ-defined depression, the overlap was significantly higher for MDD defined using at least two diagnostic codes for depression compared to MDD defined using at least one diagnostic code (Figure 3; Supplementary Table 6, including the results of comparisons).

**Figure 3:**
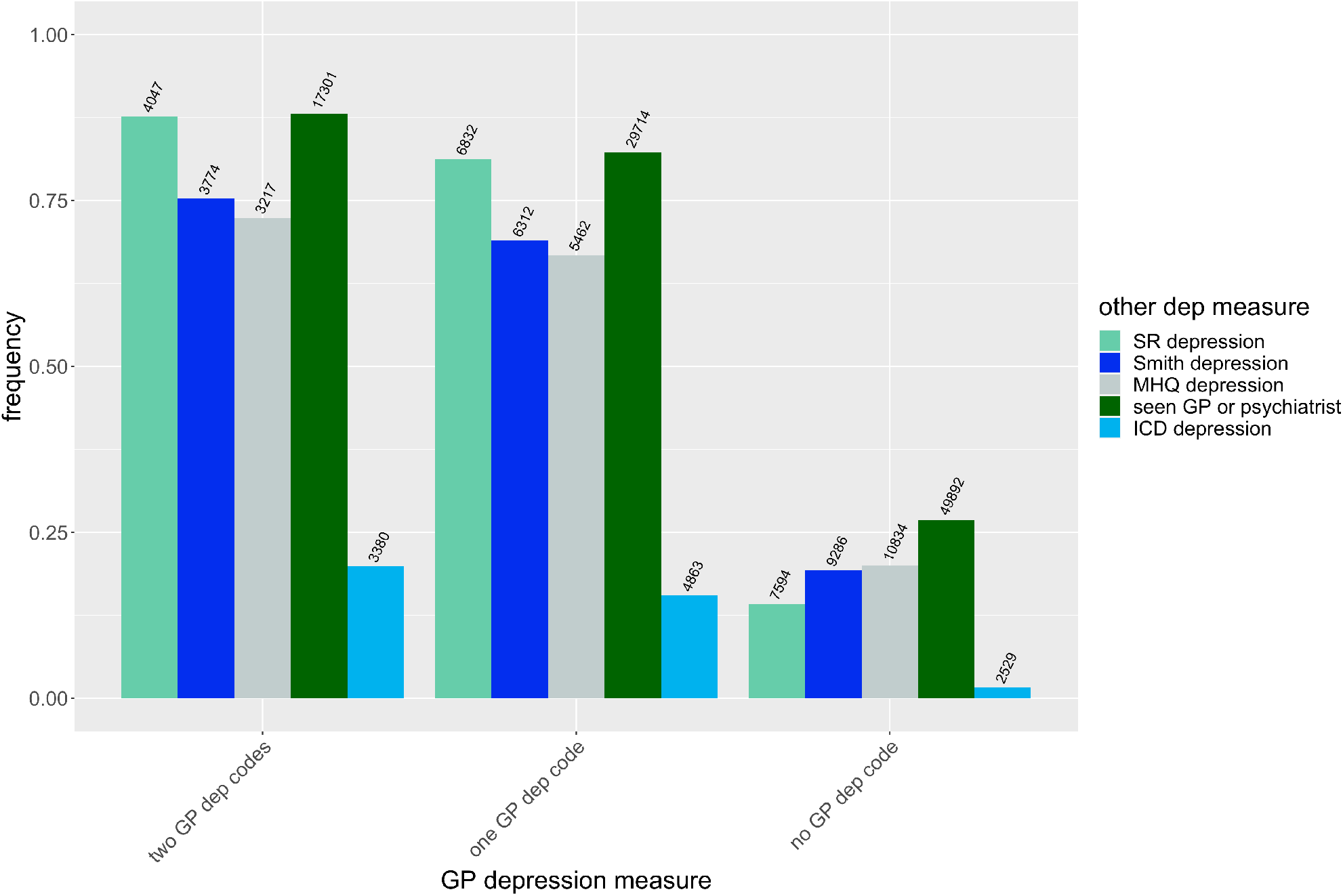
Proportion of UK Biobank participants having zero, at least one or at least two depression codes in primary care data who endorsed other measures of depression (ICD depression based on hospital records, Smith depression, self-reported (SR) depression diagnosed by a professional, MHQ (mental health questionnaire)-defined depression and help-seeking depression based on having seen a general practitioner (GP) or psychiatrist for depression-anxiety, see **Supplementary Methods**). The number of overlapping subjects is reported on top of each bar. See **Supplementary Table 6** for further details on these comparisons.

### 3.2. Clinical and socio-demographic characteristics of TRD

Participants with TRD differed from non-TRD for many clinical and sociodemographic characteristics, all indicating that TRD is a more severe and debilitating disorder (Table 1 for both cohorts, Supplementary Table 7 and Supplementary Figures 2-4 for UKB). TRD individuals were younger at first depression diagnosis as well as at first antidepressant prescription than non-TRD individuals, and they had higher BMI and higher risk of being obese than non-TRD individuals. After adjusting for potential confounders including BMI, participants with TRD did not have higher risk of type 2 diabetes and cardiovascular diseases vs. non-TRD (Table 1), though in UKB they reported a higher risk of longstanding illnesses and disabilities/infirmities vs. non-TRD (OR=2.37 [2.16-2.59], p=1.38e-75). Individuals with TRD lived in areas with higher social deprivation (Table 1), and lower SES was confirmed by other variables in UKB (education and income, Supplementary Table 7). TRD cases in UKB also showed higher levels of neuroticism and perceived loneliness, despite reporting similar rates of living alone, frequency of visits from family/friends and frequency of involvement in leisure social activities compared to non-TRD. In personality traits assessed at baseline in UKB, TRD cases reported more frequent feelings of guilt, irritability, mood swings and anxiety.

**Table 1:**
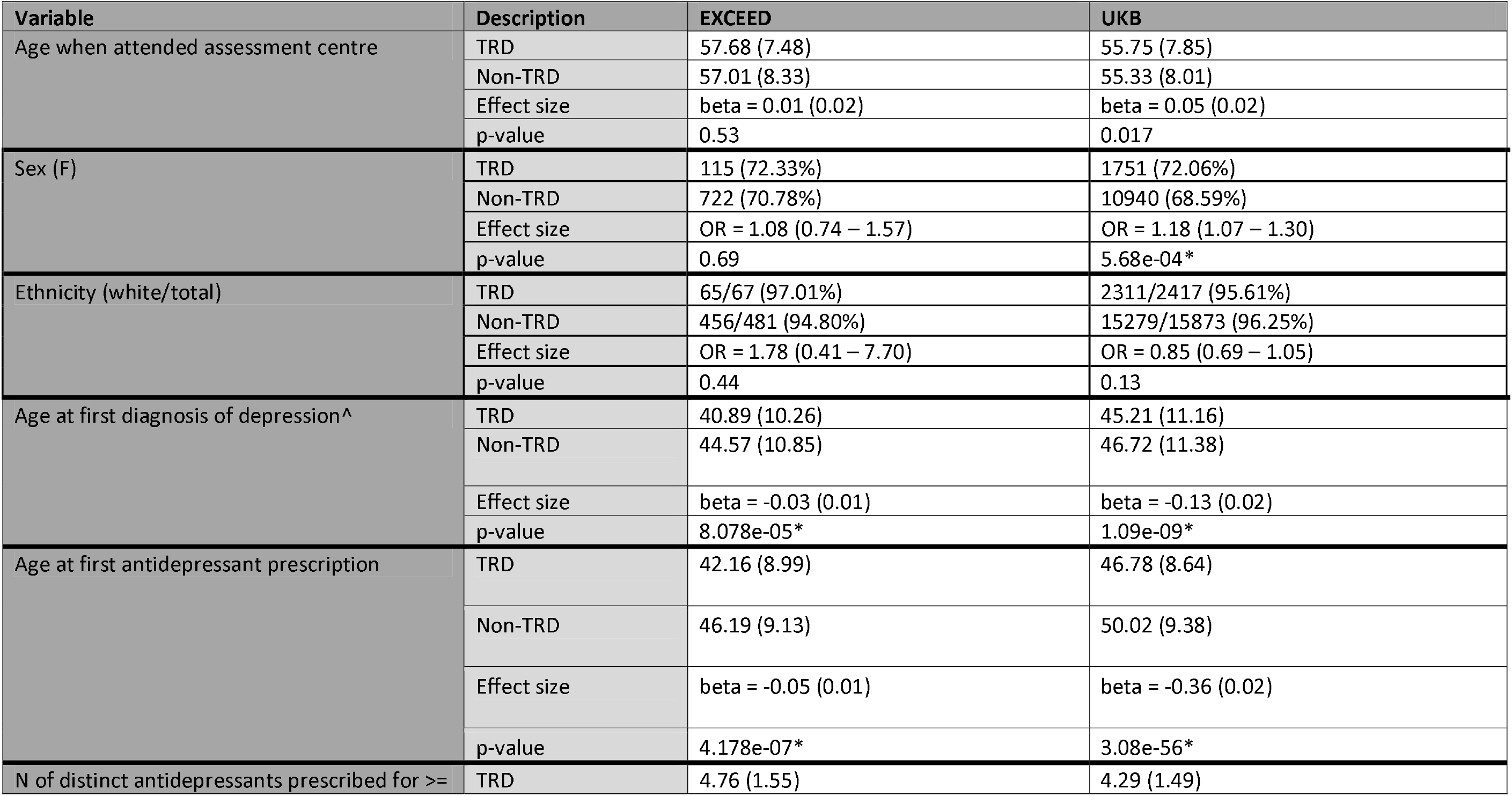
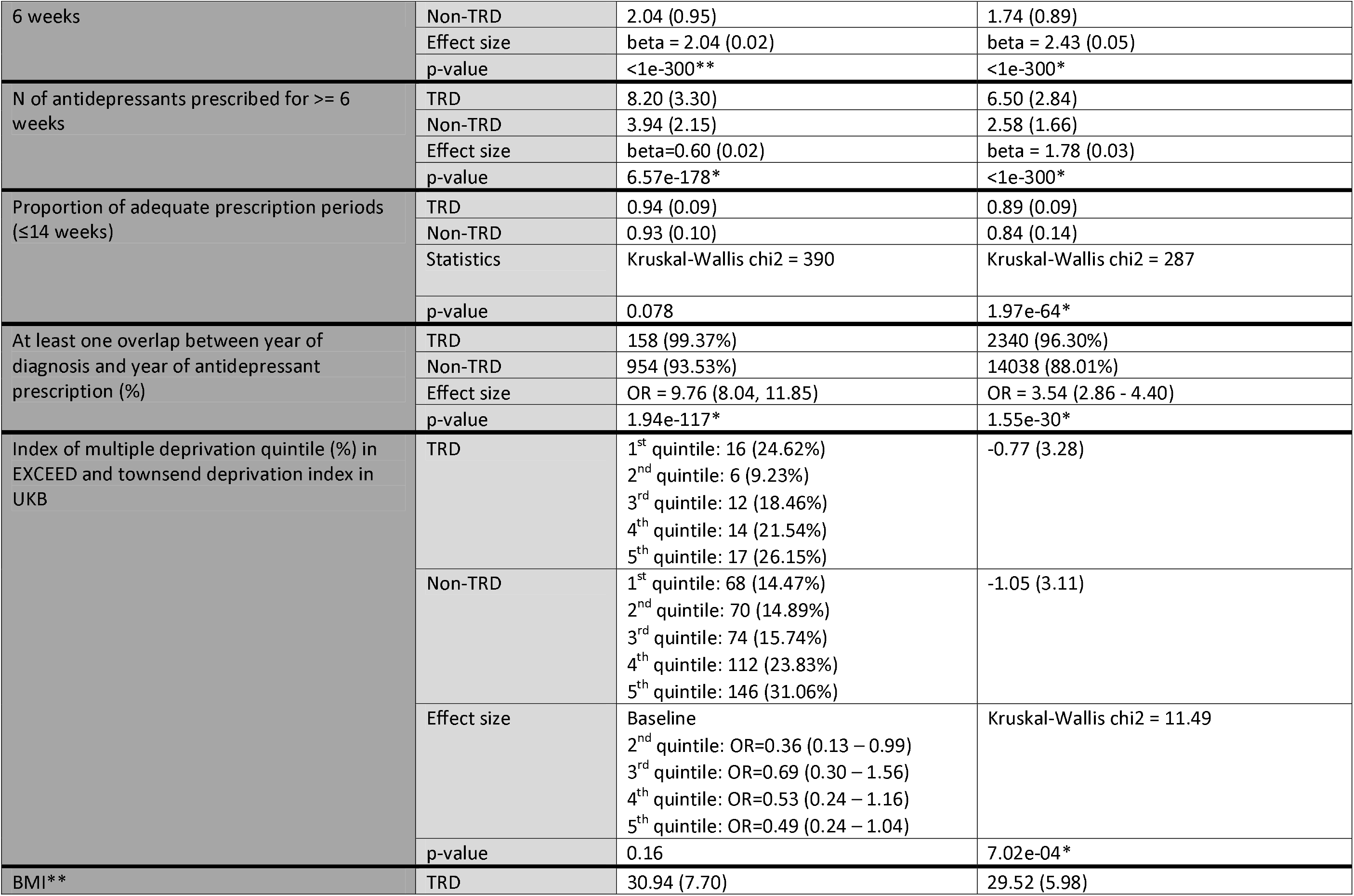
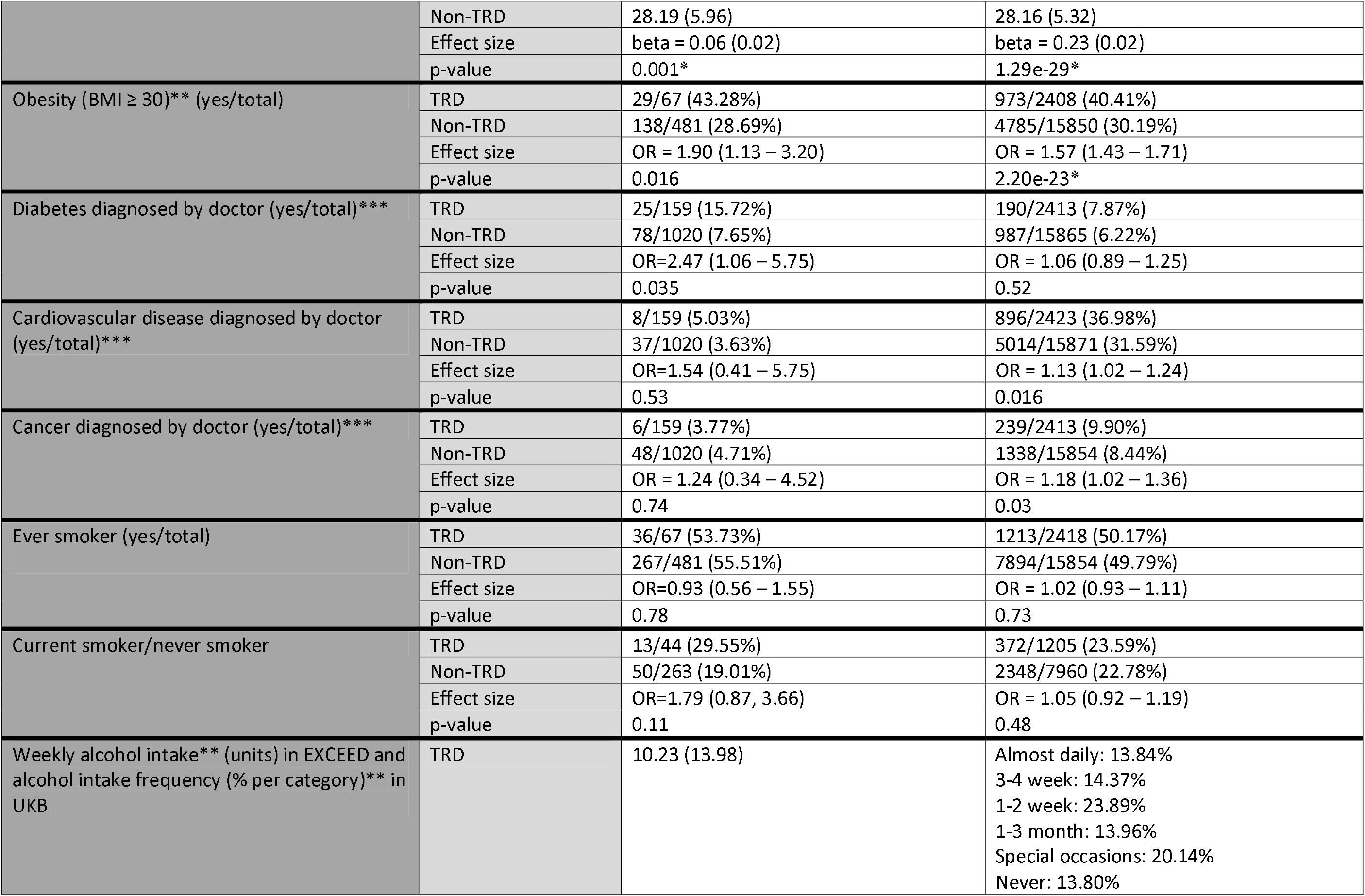
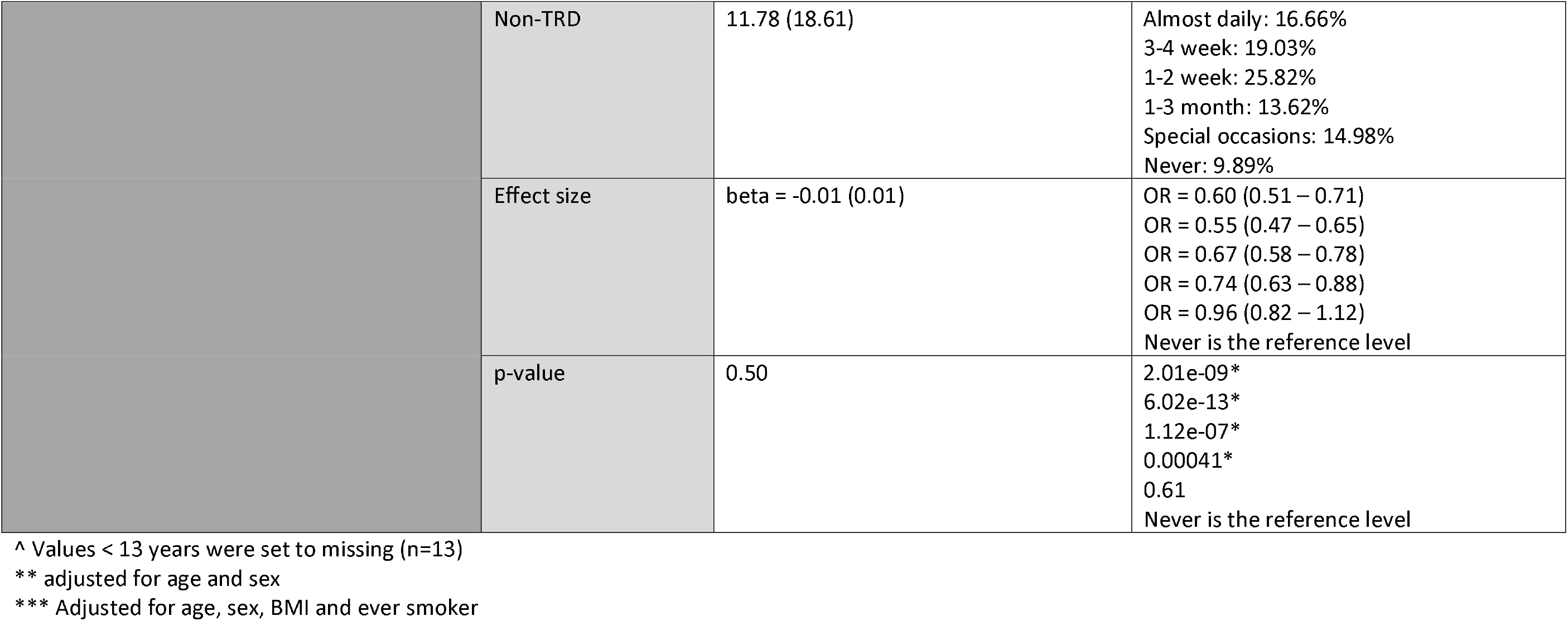
clinical and socio-demographic variables available in both cohorts and tested for association with treatment-resistant depression (TRD) vs non-TRD. Mean (standard deviation) and number (%) are reported for describing continuous and categorical variables, respectively. Analyses were logistic regression (beta (SE) or OR (95% CI) are reported for continuous and binary predictors, respectively), Kruskal-Wallis test or Cochran-Armitage trend. For categorical predictors, we compared each level to a reference level in order to provide more detailed information on the observed differences. P-values <1e-300 were reported as range. Additional variables tested in UKB are described in Supplementary Table 7. *significant p-values: alpha=1.39e-03 in UKB and alpha=0.0025 in EXCEED.

According to primary care EHR in UKB, Patients with TRD had an increased risk of comorbidity for all psychiatric disorders tested, particularly anxiety disorders (OR=1.89 [1.73-2.07], p=1.25e-45), obsessive-compulsive disorder (OR=3.03 [2.23-4.13], p=1.69e-12) and self-harm/suicidal behaviours (OR=2.03 [1.67-2.48], p=2.99e-12); Figure 4 and Supplementary Table 8. In UKB, TRD cases received psychotropic drug combinations more frequently than non-TRD patients. Antidepressant combinations were prescribed for 46% of TRD patients and 8% of non-TRD subjects (OR=5.66 [5.17-6.21], p=4.03e-289), with significant differences in the drug classes prescribed in antidepressant combinations or augmentation treatments (Supplementary Figure 5 and Supplementary Table 9). The frequency of antidepressant combinations in TRD and non-TRD was comparable in the EXCEED cohort (52.83% and 8.43%, respectively, OR=7.68 [7.42-7.94], p<2e-16). In UKB, patients with TRD vs non-TRD also had an increased probability of receiving at least one prescription of an anxiolytic/hypnotic drug (68% vs. 45%; OR=2.65 [2.42-2.91], p=2.31e-104).

**Figure 4:**
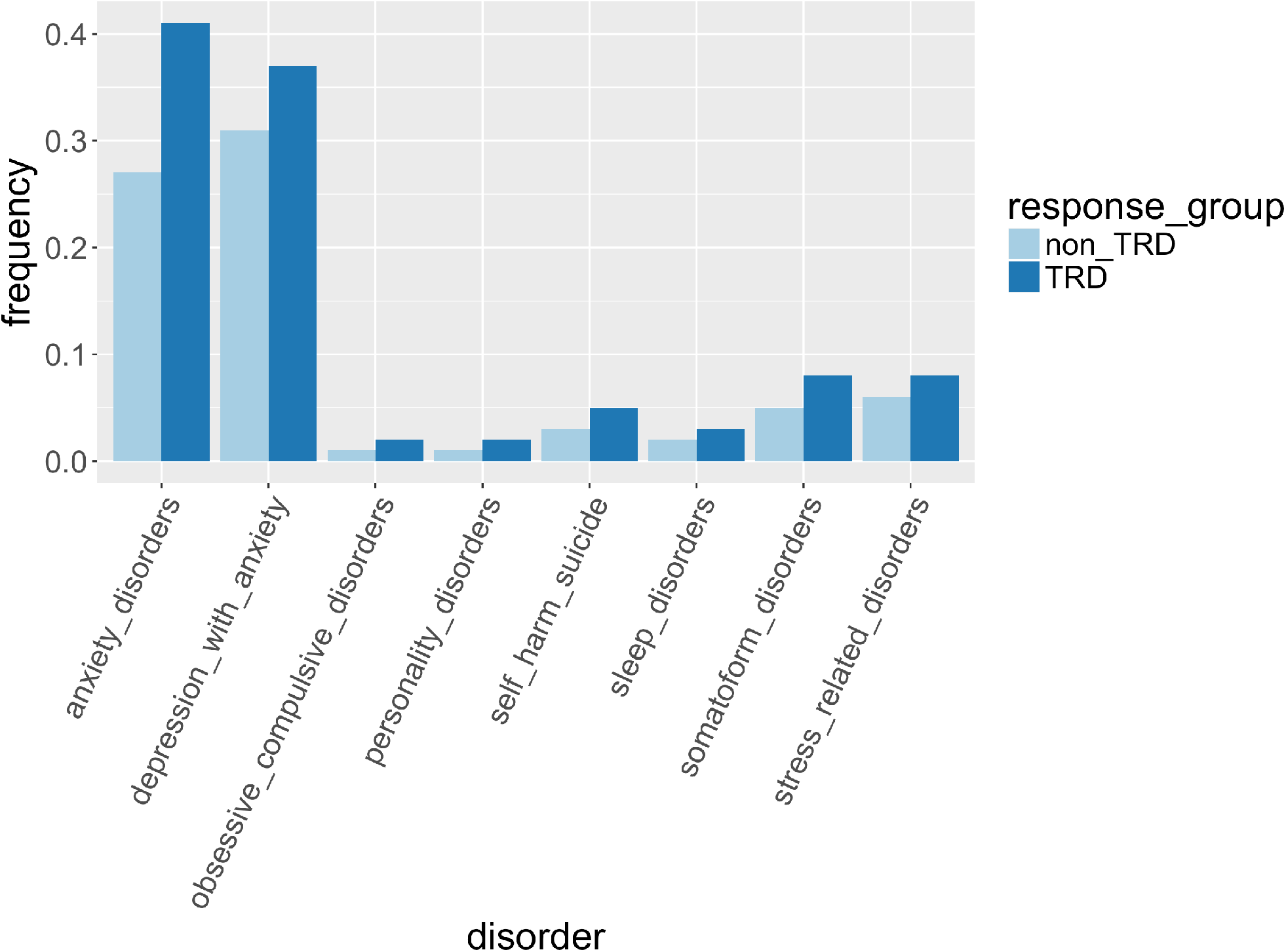
psychiatric comorbidities in patients with treatment-resistant depression (TRD) and non-TRD according to primary care records.

In UKB, a higher proportion of prescription intervals of patients classified as TRD had adequate duration compared to those of non-TRD patients, with a similar trend in EXCEED (Table 1 and Supplementary Figure 6), suggesting that lower compliance did not account for classification as TRD, though we could not determine if patients were taking the medication as prescribed.

### 3.3. Heritability of TRD in UKB

A total of 2,165 TRD and 14,207 non-TRD participants had genetic data after quality control; a further 19 and 110 individuals were excluded applying the grm-cut off. The different methods used to estimate the heritability (*h^2^_SNp_*) of TRD vs. healthy controls and non-TRD vs. healthy controls provided similar results (Table 2). On the liability scale, *h^2^_SNP_* of TRD and non-TRD were comparable, e.g. GCTB estimates were 0.25 [SE=0.04] and 0.19 [SE=0.02], respectively (z=1.17, p=0.24). The genetic correlation between TRD and non-TRD was 0.78 (SE=0.08) using LDSC, and the *h^2^_SNP_* of TRD vs. non-TRD was 0.077 (SE=0.027, p=0.004) on the observed scale (case-only comparison).

**Table 2:** SNP heritability (SNP-h2) of treatment-resistant depression (TRD) and non-TRD compared with healthy controls in UKB. After more stringent exclusion of related individuals (grm-cut off of 0.05), 2,146 cases with TRD, 14,097 cases of non-TRD and 11,188 healthy controls were included. SNP-h2 was reported according to different possible values of prevalence (*K*) in the population; the most plausible values are reported in bold. For conversion to the liability scale we used the formula reported in Supplementary Code 4.

| Method | TRD SNP-h<sup>2</sup> |  |  | Non-TRD SNP-h<sup>2</sup> |  |  |
| --- | --- | --- | --- | --- | --- | --- |
|  | Low prevalence<br>*K*=0.01 | Estimated prevalence<br>*K*=0.023 | High prevalence<br>*K*=0.03 | Low prevalence<br>*K*=0.08 | Estimated prevalence<br>*K*=0.085 | High prevalence<br>*K*=0.10 |
| GCTA | 0.2660<br>(0.0412) | **0.3306**<br>**(0.0512)** | 0.3581<br>(0.0555) | 0.2495<br>(0.0238) | **0.2563**<br>**(0.0245)** | 0.2765<br>(0.0264) |
| GCTB | 0.1979<br>(0.0333) | **0.2459**<br>**(0.0414)** | 0.2664<br>(0.0449) | 0.1877<br>(0.0185) | **0.1928**<br>**(0.0190)** | 0.2079<br>(0.0205) |
| LDSC | 0.1748<br>(0.0271) | **0.2173**<br>**(0.0337)** | 0.2354<br>(0.0365) | 0.1877<br>(0.0174) | **0.1928**<br>**(0.0178)** | 0.2080<br>(0.0193) |

For comparison, we estimated heritability for TRD reducing the stringency by additionally including those with at least one diagnostic code for depression. GCTB *h^2^_SNP_ was* not significantly different (0.22 [SE=0.04]; z=0.48, p=0.63). However, the *S* parameter suggested a different genetic architecture of these phenotypes: using at least two diagnostic codes to define MDD, both TRD and non-TRD showed *S* values significantly different from zero (p=0.003 and 0.009, respectively), but not using at least one diagnostic code (Figure 5).

**Figure 5:**
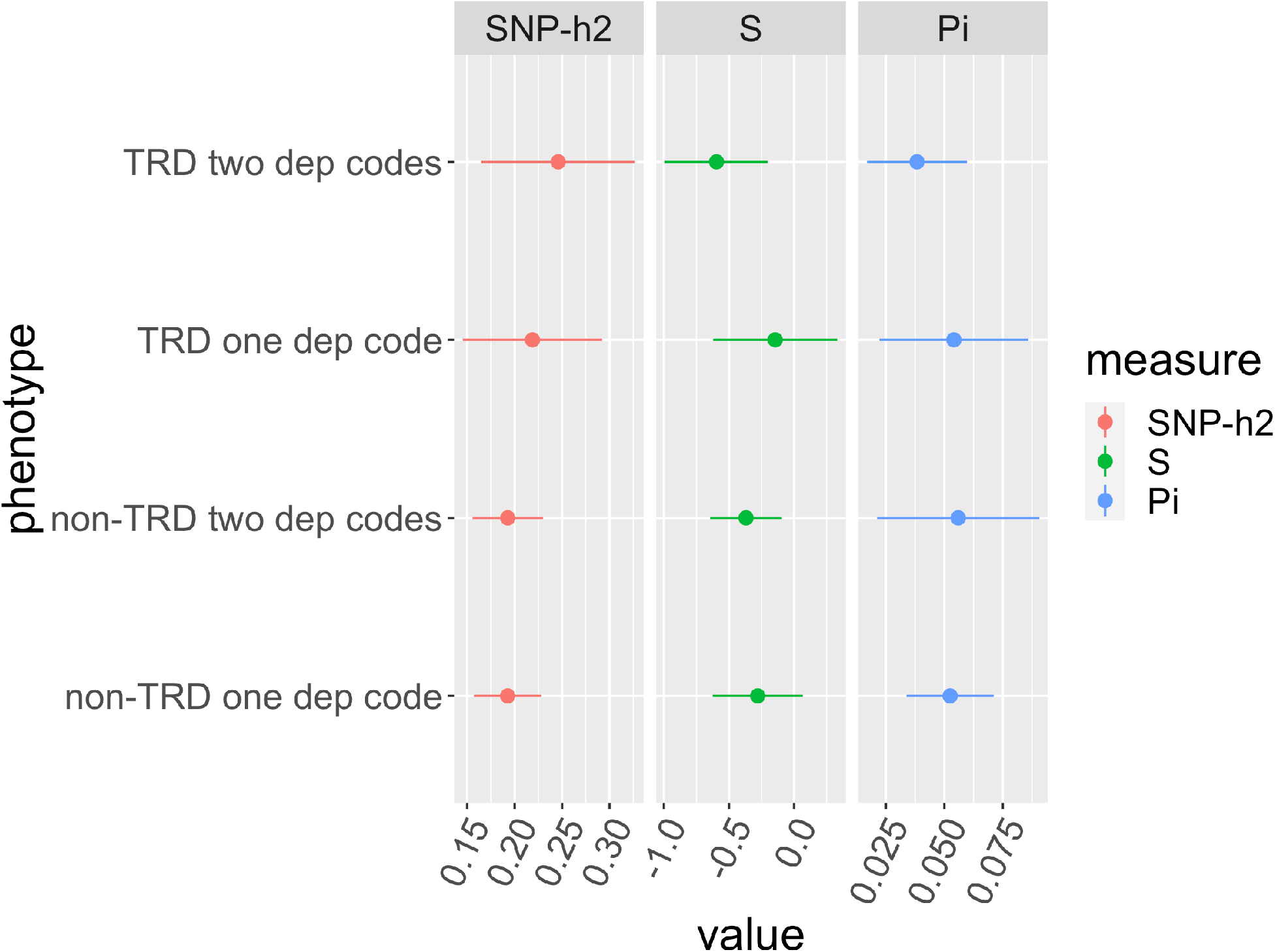
GCTB estimates of SNP-heritability (SNP-h2), negative selection (S), and polygenicity (proportion of variants with non-zero effects, Pi), for the stringent classification of TRD and non-TRD (≥ two diagnostic codes for depression), and a less stringent classification including cases with ≥ one diagnostic code for depression.

Using controls with no MDD without excluding those with other psychiatric diagnoses, the h^2^_SNP_ of non-TRD was 0.174 (SE=0.018) and the *h^2^_SNP_* of TRD was 0.233 (SE=0.040); 5 was not significantly different from zero for both phenotypes. These *h^2^_SNP_* would be 0.1360 (0.014) and 0.197 (0.034), respectively, if we consider the prevalence of having at least one code for depression instead of two as corresponding to the population prevalence of MDD from the literature.

### 3.4. Genetic correlations with other traits and PRS results in UKB

As expected, there were no genome-wide significant loci associated with TRD vs. non-TRD (variants with p < 1e-05 are in Supplementary Table 10). LDSC intercept was ^~^1 for all comparisons, suggesting no confounding factors.

Comparing TRD vs. healthy controls and non-TRD vs. healthy controls, or directly comparing TRD vs. non-TRD, we identified no significant differences in *r_g_* with other traits (Supplementary Figure 7; Supplementary Table 11). Nominally higher *r_g_* with MDD, depressive symptoms, schizophrenia, bipolar disorder, attention-deficit hyperactivity disorder (ADHD) and insomnia and lower *r_g_* with subjective well-being and childhood IQ was found for TRD vs. non-TRD (p < 0.05).

Among the PRS of psychiatric disorders, only the PRS of ADHD was significantly associated with TRD vs. non-TRD (OR=1.09 [1.04-1.14], p=4.38e-04). Among the other tested traits, the PRS of intelligence was inversely associated with TRD vs. non-TRD (OR=0.92 [0.87-0.96], p=3.08e-04), but not the PRS of childhood IQ (Supplementary Figure 8; Supplementary Table 12); results were consistent across the *P_T_* used for calculating PRS (Supplementary Figure 9).

## 4. Discussion

### 4.1. Main findings

This study demonstrated that MDD and TRD can be reliably defined using primary care records and provides the first large scale population assessment of the genetic, clinical and demographic characteristics of TRD. The prevalence of MDD found in EXCEED and UKB was 14.2% and 8.7%, respectively, which was similar to the previously reported lifetime prevalence of MDD (10.8%) (Lim et al., 2018). The significant association with MDD PRS validated our primary care-based definition of MDD, as well as the high correspondence with other measures of depression in UKB. Among individuals with MDD, those with TRD compared with non-TRD showed a number of clinical and socio-demographic features suggestive of a worse prognosis and more severe disease, such as living in areas with higher social deprivation, lower education and income, higher frequency of longstanding illnesses/disabilities/infirmities and more frequent psychiatric comorbidities. Previous studies suggested that a low SES is associated with TRD or lack of response to antidepressants (Jaffe et al., 2019; Jakubovski and Bloch, 2014).

Higher risk of chronic medical diseases as well as overweight and obesity have been previously associated with TRD (Jaffe et al., 2019; Kubitz et al., 2013; Al-Harbi, 2012; Rizvi et al., 2014), in line with the finding that higher BMI is likely to have a causal role in depression (Tyrrell et al., 2019). According to our results, BMI may be pivotal in mediating the increased rate of cardio-metabolic comorbidities in TRD. Individuals with TRD also reported less frequent moderate physical activity, which may contribute to their insufficient response to antidepressants as well as to their medical comorbidities (Blake, 2012).

In terms of personality traits, participants with TRD compared with non-TRD showed higher neuroticism, as previously described in the literature (Murphy et al., 2017), and perceived loneliness, but did not have higher probability of living alone or receiving less visits from family/friends. Higher frequency of irritability and mood swings in TRD vs. non-TRD support the hypothesis of a higher predisposition towards bipolar disorder in the former group (Murphy et al., 2017), and this hypothesis was consistent with a nominally higher *r_g_* with bipolar disorder in TRD than non-TRD, but not with our PRS results, and there was no different frequency of reported risk taking behaviours either. All the tested psychiatric comorbidities were more common in TRD than non-TRD, particularly anxiety disorders, obsessive-compulsive disorder and self-harm/suicidal behaviours, as well as psychotropic drug polypharmacotherapy, in line with previous studies (Rizvi et al., 2014; Cepeda et al., 2018; Jaffe et al., 2019; De Carlo et al., 2016; Murphy et al., 2017). The lower frequency of alcohol drinking in TRD vs. non-TRD does not imply a lower risk of alcohol use disorders, and it does not reflect the amount of alcohol drunk per drinking episode; in the context of our study, this finding may be related to the lower SES of participants with TRD than non-TRD (Galea et al., 2007; Casswell et al., 2003; Beard et al., 2019).

Our results suggested that TRD has a similar *h^2^_SNP_* compared with non-TRD and both these groups may have a different genetic architecture when defined in subjects having at least one diagnostic code for depression rather than at least two, but non-significantly different *h^2^_SNP_*. In participants having at least two diagnostic codes for depression compared to those having at least one we found indeed a negative GCTB *S* parameter, that was proposed as a marker of negative selection (Zeng et al., 2018). *S* was not significantly different from zero when considering controls screened for MDD but not for other psychiatric disorders, while *h^2^_SNP_* was similar. Compared to a previous GWAS of TRD vs. healthy controls in unrelated individuals (Li et al., 2016), our *h^2^_SNP_* estimate was similar according to both GCTB (z=1.16, p=0.25) and LDSC (z=0.79, p=0.43). Contrary to this previous study, our *h^2^_SNP_* of TRD vs. non-TRD was significantly different from zero.

Genetic correlations with other traits and PRS analyses supported that TRD shares genetic predisposition with ADHD compared to non-TRD. Undetected ADHD was associated with the risk of SSRI-failure and a higher number of previous medications in depressed individuals (Sternat et al., 2018). Interestingly, 17%-22% of adults attending psychiatric outpatient clinics for conditions other than ADHD were found to suffer from this disorder; however, fewer than 20% of adults with ADHD are diagnosed and/or treated by psychiatrists (Ginsberg et al., 2014). Therefore, undiagnosed ADHD or a past diagnosis of the disease should be assessed in patients with TRD. In our sample, ADHD diagnosis was difficult to assess, as it could not be reliably assessed in primary care data (prevalence < 0.01%), probably due to the lack of registration of diagnoses received during childhood, lower awareness of the manifestations of ADHD and underdiagnosis in the past as well as underdiagnosis in adults (Ginsberg et al., 2014; Polyzoi et al., 2018). Although not included in our main analyses due to availability in a subset of the UKB only, the OR of self-reported ADHD diagnosis received by a professional was 6.65 (95% CI 2.04-21.07, p=8.12e-04) in TRD compared with non-TRD groups (7 on a total of 581 and 8 on a total of 4373 subjects, respectively).

PRS analyses suggested that the PRS of intelligence was inversely associated with TRD compared to non-TRD. Discordant results have been previously reported about the *r_g_* between Intelligence and MDD/depression (Howard et al., 2019; Wray et al., 2018; Krapohl et al., 2018; Sniekers et al., 2017). To the best of our knowledge, no previous study demonstrated that the PRS of intelligence may be negatively associated with TRD compared with non-TRD, but lower intelligence was reported to be a predictor of poor antidepressant response (Fournier et al., 2009). This finding may be related to SES differences, but our *r_g_* results for educational attainment did not support a difference between TRD and non-TRD, therefore the intelligence PRS result should be interpreted cautiously.

### 4.2. Limitations

EHR used to define TRD and non-TRD groups do not necessarily reflect complete information regarding antidepressant prescription, particularly for prescriptions issued before 1990s; therefore, a part of the prescriptions as well as diagnoses may be missing for the included participants. Compliance to prescribed drugs may be as low as 50% in outpatient settings (Sansone and Sansone, 2012), therefore our TRD rate may be overestimated; however, it is in the range of previous literature estimates and we considered the time between consecutive prescriptions to assess this issue. The lack of a standardised diagnostic assessment together with the observed variety of diagnoses may have led to the inclusion of cases with depressive disorders other than MDD, however the overlap with other measures of MDD and the genetic overlap with MDD suggest that this issue was mild at most. We did not have a direct measure of treatment response, but we assumed that a switch to a different antidepressant was indicative of lack of efficacy when there the first one was prescribed for at least six weeks, since a switch due to side effects would probably happen earlier (Wigmore et al., 2020). Moreover, switching is the most common strategy for TRD management (MacQueen et al., 2017). Prescribed daily medication dose was not available, therefore was not used for the definition of TRD, and subtherapeutic doses may have inflated the observed TRD rate. Primary care data were available only in about half of UKB participants, therefore there was limited overlap with variables assessed in other subsets of the sample, such as the MHQ. The low prevalence of some disorders in primary care data, such as personality disorders, is likely caused by the lack of training of general practitioners (GP) for assessing these diagnoses, as shown by the low agreement between GP diagnosis and structured interviews, and the previously reported low prevalence in primary care (Moran et al., 2001; Wlodarczyk et al., 2018).

Regarding the genetic part of the study, we had inadequate power to identify variants associated with TRD versus non-TRD at the genome-wide level (Visscher et al., 2017) or to use the generated summary statistics to create a PRS of TRD in other samples. Though there was a nominally higher *r_g_* with insomnia for TRD vs. non-TRD, we did not test the PRS of insomnia because the two main GWAS available were either based on UKB (Hammerschlag et al., 2017) or about the 30% of the sample was represented by UKB participants (The 23andMe Research Team et al., 2019), and this could bias the results. The PRS of subjective well-being was obtained in a sample with a partial overlap with the UKB cohort of this study (1.6% of the sample (Okbay et al., 2016)).

### 4.3. Conclusions

This study demonstrated that MDD can be reliably defined according to EHR of primary care events as it shows good comparability with other depression measures. These data can be also used to identify patients with TRD and study their phenotypic and genetic characteristics compared with non-TRD. Our results suggested that TRD has partially distinct genetic characteristics compared with non-TRD and a number of socio-demographic and clinical characteristics suggestive of a higher disease severity and worse prognosis. These included variables that may be helpful to identify patients at risk of TRD who should be considered for referral to secondary care. Social policies should promote awareness of the factors associated with TRD and its negative consequences on health, as well as aim to reduce inequalities related to SES as these are likely to impact on the risk of TRD among people with depression.

## Data Availability

UK Biobank resource is available for researchers with an approved application (https://www.ukbiobank.ac.uk/wp-content/uploads/2014/06/1000-Naomi-Allen-10am-data-and-access.pdf). The code used to generate the results of this work has been included as supplementary material. 
For information on the EXCEED cohort see: https://www.hra.nhs.uk/planning-and-improving-research/application-summaries/research-summaries/extended-cohort-for-e-health-environment-and-dna-exceed-study/

## Acknowledgements

The authors acknowledge use of the research computing facility at King’s College London, Rosalind (https://rosalind.kcl.ac.uk), which is delivered in partnership with the NIHR Biomedical Research Centres at South London & Maudsley and Guy’s & St. Thomas’ NHS Foundation Trusts, and part-funded by capital equipment grants from the Maudsley Charity (award 980) and Guy’s & St. Thomas’ Charity (TR130505). This research has been conducted using the UK Biobank Resource under Application Number 56514 “Stratification of health outcomes in mood disorders”.

The EXCEED study gratefully acknowledges the support of all participants and staff who have contributed to the study. This research used the ALICE High Performance Computing Facility at the University of Leicester.

